# Feasibility and safety of shortened hypofractionated high dose palliative lung radiotherapy – A retrospective planning study

**DOI:** 10.1101/2021.11.10.21265927

**Authors:** Matthew Jones, Jane Rogers, Raj Kumar Shrimali, Jo Hamilton, Senthil Athmanathan, Bleddyn Jones

## Abstract

**Objective:** Assess the safety and feasibility of shortened hypo-fractionated high dose palliative lung radiotherapy in a retrospective planning study.

**Methods:** Fifteen palliative lung radiotherapy patients previously treated with the standard 36 Gy in 12 fractions (12F) schedule were non-randomly selected to achieve a representative distribution of tumour sizes, volumes, locations and staging. Plans were produced using 30 Gy in 5 fractions (5F) and 30 Gy in 6 fractions (6F) using a 6MV FFF co-planar VMAT technique. These plans were optimised to meet dose constraints for both PTVs and OARs where established OAR constraints were expressed as BED. The potential safety of the delivery of these plans was assessed using these BEDs and also with reductions of 10% and 20% to account for effects of chemotherapy or surgery.

**Results:** For all 5F and 6F plans the mandatory constraints using the full BED were met, as with all 6F plans with 10% BED reductions, but reduced to 6 patients using 5F. Three of 15 5F and 6 of 15 6F plans met the 20% BED reductions.

**Conclusion:** It is potentially safe and feasible to deliver high dose palliative radiotherapy using the 5F or 6F regimes described, when carefully planned to comparable OAR BEDs as for 12F standard. It appears that the toxicity from either of these regimes should be within acceptable limits provided that the dose constraints described can be adhered to. A Phase II study would be required to fully assess the safety and feasibility. The outcomes from such a study could potentially reduce the number of patient hospital visits for radiotherapy, thus benefiting the patient and the clinical service by optimising resource utilisation.

**Advances in Knowledge:** Shortened hypo-fractionated high dose palliative lung radiotherapy is technically feasible provided OAR constraints are respected.

## Introduction

Non-Small Cell Lung Cancer (NSCLC) is a leading cause of cancer related deaths in the UK and worldwide (1–4). Over three-quarter of all patients with NSCLC patients present with late stage (stage III and IV) disease and about half present with metastatic (stage IV) disease (3–5). About 48,000 patients in the UK (39,000 patients in England) were diagnosed with lung cancer in 2017, and about half of these patients presented with stage IV disease (3,4).

The treatment for stage IV lung cancer is aimed at maintaining or improving quality of life (by controlling symptoms) and prolonging survival. These patients are not candidates for curative therapy due to presence of metastases, tumour volume, poor ECOG performance status or comorbidities (6), and have a 1-year survival of under 20% and a 5-year survival of about 3% (4). Systemic therapy is the primary treatment for patients with advanced NSCLC. Advances in chemotherapy, targeted agents and immunotherapy has resulted in survival improvement (7–14). This has led to a significant expansion in use of systemic treatment options in the last 15 years, including targeted therapy using tyrosine kinase inhibitors (TKIs), immunotherapy, chemotherapy and combination treatments (chemotherapy and immunotherapy) (14). However, radiotherapy is also still used in about 30% of patients with stage IV NSCLC, as required on a case-by-case basis, for local control and symptomatic metastases from advanced non-small cell lung cancer (5). Palliative thoracic radiotherapy is also used for patients who are unresponsive to systemic therapy, disease progression after initial response, upon relapse after systemic therapy and those who are not suitable for systemic therapy (15).

A phase III randomised controlled trial (RCT) reported by Macbeth *et al*., and a subsequent systematic review by Fairchild *et al*., have shown that, in patients with good performance status, higher dose regimes equal to or greater than 30 Gy in 10 fractions (3 Gy per fraction) were associated with improvements in total symptom score and survival (but at the cost of increased side effects, such as radiation oesophagitis) (16,17). Other studies did not confirm this (16,18–23). A Cochrane review update on palliative thoracic radiotherapy, published in 2015, looked at 3576 patients treated in 14 randomised controlled trials, using a total of 19 different dose fractionation regimes, and found large differences in patient characteristics, performance status and outcome measures (24). Higher doses or more fractionated palliative radiotherapy regimens were not shown to provide better or more durable palliation or to prolong survival (24). More recently, other studies have suggested that higher doses of palliative thoracic radiotherapy were associated with improved survival (25–27). High dose palliative radiotherapy using 30-39 Gy in 10-13 fractions has been recommended, in a statement by the Royal College of Radiologists, for patients with better performance status (0-2) who may be suitable candidates for multiple lines of systemic therapy (28).

Clinically relevant organs at risk (OARs) from the literature in this setting include oesophagus, normal lung tissue and the spinal cord. Within the Cochrane review, radiation related oesophagitis was reported in 8 studies (1301 patients) either as patient-reported toxicity or physician assessed toxicity. The mean rate of grade 3-4 oesophagitis was reported as 25.7% in the intervention groups and 22.3% within the control groups. Radiation pneumonitis was not graded or reported in most trials, with the mean rate of 3.9% in the control groups and 2.4% in the intervention group (24). Spinal cord remains an organ of concern because of the rare but serious late complication of radiation myelopathy (RM), that was reported in 3 patients treated in the RCT by the Medical Research Council Lung Cancer Working Party (MRC-LCWP) in 1996 (16). At the time, the pooled data were analysed on 1048 patients from three randomized trials conducted by the MRC-LCWP (29). Although patients in this analysis were treated with 7 different palliative lung RT regimens, RM was reported in a small number of patients treated with 17 Gy in 2 fractions (1 week apart) and 39 Gy in 13 fractions (5 fractions/week). Cumulative risks of RM by 2 years were estimated at were 2.2% for the 17 Gy group, 2.5% for the 39 Gy group (29). The 30-day mortality was found to be 9% in a large retrospective single-centre study and about 14% in a large population-based study (25,30). The median survival after palliative RT ranged from 6.2 to 9 months and 1 year survival was reported as 39% (16,24).

Most of the treatment strategies are based on randomised evidence generated in the 1990s when radiotherapy planning was comparatively simple (parallel-pair or 3-dimensional conformal radiotherapy) and treatment delivery less precise. The dose-fractionation used in trials from that era remain the basis for current palliative lung RT practice (28). Modern radiotherapy techniques typically comprise of highly conformal radiotherapy using intensity modulated radiotherapy (IMRT) or volumetric modulated arc therapy (VMAT) aided by volumetric verification using cone beam computed tomography (CBCT) to ensure that the planned treatment is accurately delivered to the target volume. The desired dose distribution is achieved in the planning target volume (PTV), keeping the doses to normal organs within tolerance (as defined by dose constraints).

We hypothesize that it may be possible to safely treat patients with an adequate dose of palliative radiation comparable to the current standard of care (36 Gy in 12 fractions), by using 30 Gy in 5 or 6 fractions, given on alternate days, provided that normal tissue dose constraints are met. The associated reduction in fractions would reduce the number of patient visits to the radiotherapy department and the overall duration of radiotherapy. Besides the obvious cost saving, more radiotherapy treatment capacity would follow.

In this retrospective treatment planning study, we aim to assess the feasibility and theoretical safety of reducing the duration and number of fractions for palliative thoracic radiotherapy, based on established dose constraints and computed biological equivalent doses (BED). The safety would be assessed with a particular focus on the planned dose distributions to the OARs (spinal cord, oesophagus and normal lung tissue), as compared against palliative radiotherapy plans for the current standard of 36 Gy in 12 fractions.

## Materials and Methods

Fifteen patients previously treated at our centre with 36 Gy in 12 fractions were included within the planning study. This cohort of patients was non-randomly selected to achieve a reasonable distribution of tumour sizes, volumes, anatomical location and staging from all patients previously treated with this fractionation.

For each patient, the following OARs were outlined if they were not present in the original treatment plan: spinal cord, heart, lungs, oesophagus, great vessels, brachial plexus, trachea, proximal bronchial tree and chest wall. All organs were outlined following the current UK SABR consortium guidelines (31) and a 5 mm isotropic margin was added to the spinal cord to create a planning risk volume (PRV). A 5 mm isotropic margin was added to the gross tumour volume (GTV) to create the clinical target volume (CTV) and a 1 cm isotropic margin was added to this to create the planning target volume (PTV) in line with standard practice at our centre for palliative lung patients.

A 6MV co-planar VMAT technique was used to produce the 36 Gy in 12 fractions (12F) plans for all patients and plans were optimised with the aim of covering the PTV with at least 95% of the prescribed dose. No further dose constraints were used for either target volumes or OARs in line with standard practice at our centre for palliative lung patients. For the 30 Gy in 5 fractions (5F) and 30 Gy in 6 fractions (6F) plans, a 6MV VMAT technique was used. To minimise the treatment time, a flattening filter free beam was used instead of a conventional flattened beam for both 5F and 6F plans. Both sets of plans were optimised to meet the clinical goals for both target volumes and OARs given in Table 1. The RayStation treatment planning system (RaySearch Laboratories AB, Stockholm, Sweden) was used for all plans with an Elekta (Elekta AB, Stockholm, Sweden) Agility beam model.

**Table 1:** Dose constraints and alpha beta ratios used for all OARs within the study.

| Organ | Comparator | Dose in Gy | Volume in cc | Suggested Priority | Source | $\alpha/\beta$ |
| --- | --- | --- | --- | --- | --- | --- |
| Ipsilateral Brachial Plexus | At most | 30 | 3 | Optimal | (32) | 3 |
| Ipsilateral Brachial Plexus | At most | 32 | 0.1 | Mandatory | (32) | 3 |
| Heart / Pericardium | At most | 32 | 15 | Mandatory | (32) | 3 |
| Heart / Pericardium | At most | 105% Presc | 0.1 | Mandatory | (32) | 3 |
| Oesophagus - early | At most | 34 | 0.5 | Mandatory | (31) | 10 |
| Oesophagus - early | At most | 32 | 0.5 | Optimal | (31) | 10 |
| Oesophagus - early | At most | 27.5 | 5 | Optimal | (32) | 10 |
| Oesophagus - late | At most | 34 | 0.5 | Mandatory | (31) | 3 |
| Oesophagus - late | At most | 32 | 0.5 | Optimal | (31) | 3 |
| Oesophagus - late | At most | 27.5 | 5 | Optimal | (32) | 3 |
| Spinal Canal PRV | At most | 30 | 0.1 | Mandatory | (31) | 2 |
| Spinal Canal | At most | 23 | 0.1 | Optimal | (31) | 2 |
| Trachea and Bronchus - early | At most | 35 | 0.5 | Mandatory | (31) | 10 |
| Trachea and Bronchus - early | At most | 32 | 0.5 | Optimal | (31) | 10 |
| Trachea and Bronchus - late | At most | 35 | 0.5 | Mandatory | (31) | 3 |
| Trachea and Bronchus - late | At most | 32 | 0.5 | Optimal | (31) | 3 |
| Lung - PTV | At most | V14.7Gy | 20% | Optimal | (*) | 3 |
| Lung - PTV | At most | V20Gy | 20% |  | (*) | 3 |
| Chest Wall | At most | 32 | 30 | Optimal | (31) | 3 |
| Great Vessels | At most | 47 | 10 | Mandatory | (32) | 3 |

Dose constraints for the 5F and 6F plans were taken from multiple sources which included the UK SABR Consortium Guidelines (31), RCR Guidelines for reduced fractionation during the COVID-19 pandemic (32) and local protocols, or derived from our local dose constraints for radical lung treatments derived from the SOCCAR trial protocol (33).

The BED is defined as:

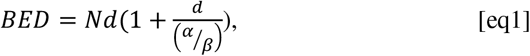

Where, N is the number of fractions, d the dose per fraction and α/β is the biological parameter that influences fractionation sensitivity. There are many applications of BED in radiotherapy (34–36). The α/β ratios used are 2 Gy for RM and 3 Gy for other normal tissue late effects, and 7 or 10 Gy for squamous cell cancers.

The EQD-2, the number of 2 Gy fractions which provide the same BED, is obtained from the BED as:

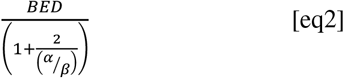

The BED for each OAR was calculated for all 5F, 6F and 12F plans using the values of α/β given in Table 1. The absolute and percentage difference in BED was compared between all three fractionation regimes. OARs highlighted in yellow were regarded as the most important regarding safety. The TCP and NTCP methods favoured by some authors are not used since they involve further considerable assumptions e.g., tumour cell density and normal tissue structural complexity and are not regarded as reliable for individual predictions.

For this reason, the three fractionation regimes are compared for tumour control by BED estimations only (further details below), where a α/β ratios of 10 Gy and 7 Gy are used, as in table 2. Higher values of α/β may occur in very poorly differentiated tumours but further increments in α/β beyond 7-10 Gy have little influence on isoeffective dose estimations.

**Table 2:** BED and EQD-2 of prescribed doses for tumour control

|  | **36 Gy in 12#** | **30 Gy in 6#** | **30 Gy in 5#** |
| --- | --- | --- | --- |
| **BED ($\alpha/\beta=10$ Gy)** | 46.8 | 45 | 48 |
| **EQD-2 ($\alpha/\beta=10$ Gy)** | 39 | 37.5 | 40 |
| **BED ($\alpha/\beta=7$ Gy)** | 51.4 | 51.4 | 55.7 |
| **EQD-2 ($\alpha/\beta=7$ Gy)** | 40 | 40 | 43.3 |

It can be seen from Table 2 that the three schedules have broadly similar tumour BEDs and EQD-2, although the highest values are obtained with the five-fraction schedule, but which can only be permitted if the normal tissue constraints are met. No repopulation factor for tumour cell proliferation is used here as this is known to be minimal below 21 days in squamous cell cancers (37), which applies to all three schedules. The three most complex plans as determined by having the highest calculated modulation factors (M) were delivered to a Delta4 phantom (Scandidos, Uppsala, Sweden) and evaluated using gamma analysis using local clinical tolerances. The modulation factor is a local measure of plan complexity which represents the monitor units per Gray required to deliver the prescribed dose for a given treatment plan and is therefore defined as:

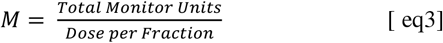

It is known that the use of most forms of chemotherapy given either simultaneously or sequentially can reduce the BED constraints in most organs at risk and especially the nervous system by causing sub-clinical tissue vascular damage that will reduce radiation tolerance (38). The tolerance changes used in clinical practice are generally a BED reduction of around 10%. Furthermore, a history of surgery affecting the OAR, major trauma or multiple chemotherapy courses and extreme age should reduce the BED constraints even further, by about a total of 20% in the worst-case scenario (39).

The reduced constraints are displayed by lines representing full constraint and minus 10% and 20% of BED. These ranges of reductions have been favoured in difficult clinical situations where the risk of RM is high, as in radiotherapy retreatments which include the spinal cord (40,41). The range of 10-20% reduction of BED is consistent with pragmatic choices of fractionation commonly used to respect spinal cord tolerance depending on clinical circumstances and in different Cancer Centres. For example, table 3 shows an 8 to 20% change in BED for these schedules.

**Table 3:** Changes in BED and EQD-2 for different fractionation schedules with 50 Gy in 25 fractions representing spinal cord tolerance associated with a risk of 0.083% in patients with no adverse histories [Wooley et al.].

|  | **50 Gy in 25#** | **46 Gy in 23#** | **45 Gy in 25#** | **40 Gy in 20#** |
| --- | --- | --- | --- | --- |
| **BED ($Gy_{[2]}$)** | 100 | 92<br>(-8%) | 85.5<br>(-14.5%) | 80<br>(-20%) |
| **EQD-2\*(Gy)** | 50 | 46<br>(-8%) | 42<br>(-14.5%) | 40<br>(-20%) |

Similar percentage BED tolerance reductions are used for all other normal tissue late reactions, but with use of α/β=3 Gy, which makes little difference to the percentage changes.

## Results

Figure 1 shows the BEDs to the OARs (highlighted in Table 1) considered to be the most important for patient safety. BEDs were derived from the radiotherapy plans for each of the 15 patients (listed 1 – 15), with fractionations of 36 Gy in 12F, 30 Gy in 5F and 30 Gy in 6F (shown as indicated by striped green, solid blue, and hashed red, respectively). The relevant mandatory (thick purple) and optimal (thin orange) constraints are indicated in terms of the BED converted from the dose constraint defined for 30 Gy in 5F. Also shown in (d) is the spinal canal PRV relative to the more conservative standard BED constraint of 100 Gy _[2]_, the subscript in square parentheses denoting that an α/β of 2 Gy was used.

**Figure 1:**
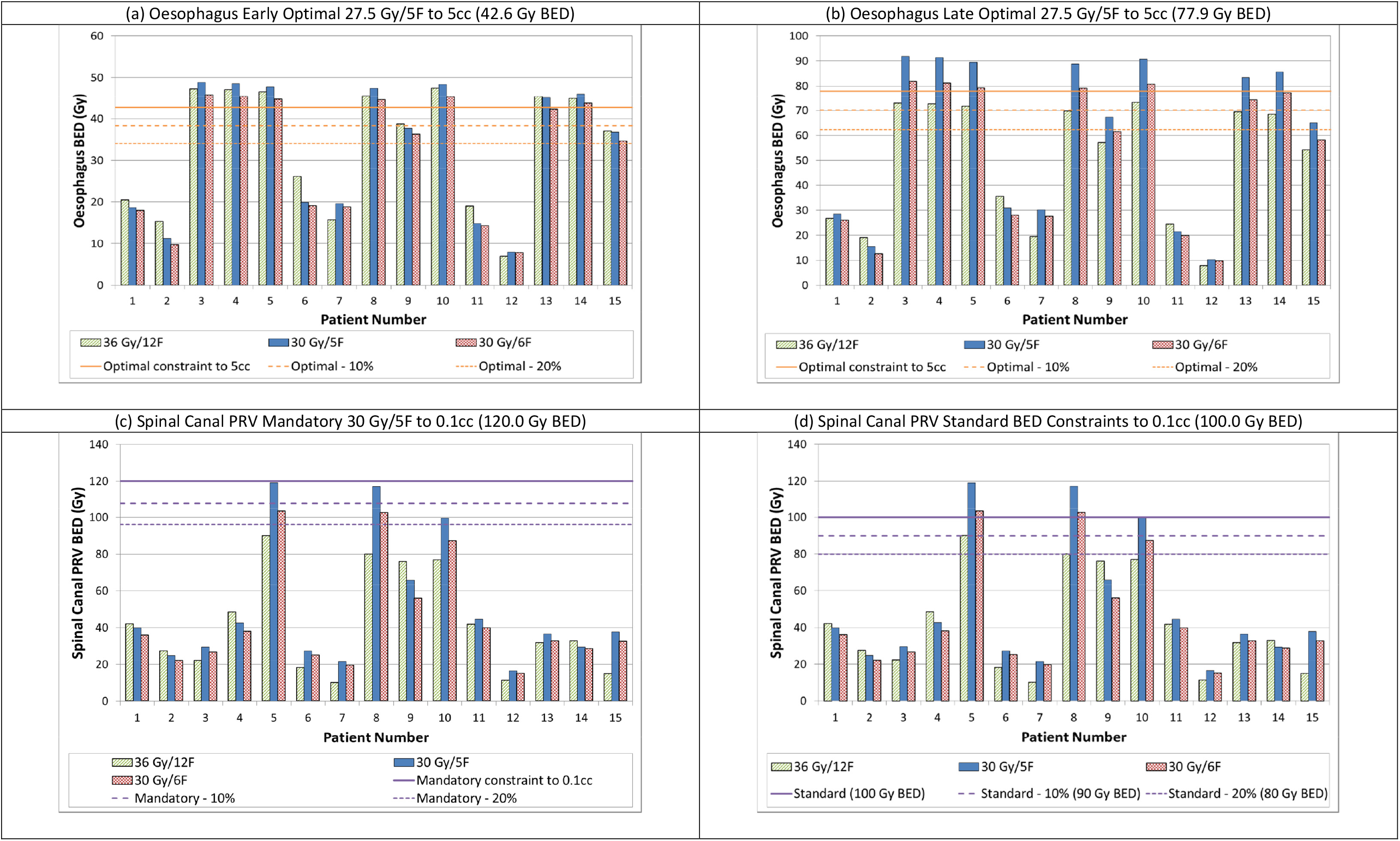
Biologically Equivalent Doses (BEDs) to OARs highlighted in Table 1 as most important for safety, for 15 patients (numbered 1 – 15). Fractionations of 36 Gy in 12F, 30 Gy in 5F and 30 Gy in 6F are shown as indicated by striped green, solid blue, and hashed red, respectively. The relevant mandatory (thick purple) and optimal (thin orange) constraints are indicated in terms of the BED converted from the dose constraint defined for 36 Gy/12F. Long and short dashed lines indicate the estimated relevant constraints with chemotherapy (standard constraint – 10%) and with chemotherapy plus surgery (standard constraint – 20%) respectively. Data are shown for (a) Oesophagus early optimal constraint 27.5 Gy/5F to 5cc (42.6 Gy BED), (b) Oesophagus late optimal constraint 27.5 Gy/5F to 5cc (77.9 Gy BED), (c) Spinal canal PRV mandatory constraint 30 Gy/5F to 0.1cc (120.0 Gy BED), (d) Spinal canal PRV with standard (i.e. more conservative) BED constraint taken from Table 3 to 0.1cc (100.0 Gy BED).

Long and short dashed lines indicate the estimated relevant reduced constraints because of addition of chemotherapy (standard constraint – 10%) and further reduction of constraints because of greater damage caused by multiple chemotherapy lines of chemotherapy plus surgery or significant trauma (standard constraint – 20%), respectively.

Figure 2 shows the Lung – PTV percentage volumes receiving (a) 14.7 Gy and (b) 20 Gy for patients 1-15 and planned for the 3 fractionations and with constraints indicated as in Figure 1. Figure 3 shows the BEDs for the remaining OARs from Table 1 with long and short dashed lines indicating the additional constraints by chemotherapy and medical history as per Figure 1. PTV constraints as detailed in Table 1 (in Methods) were all met in all of the plans.

**Figure 2:**
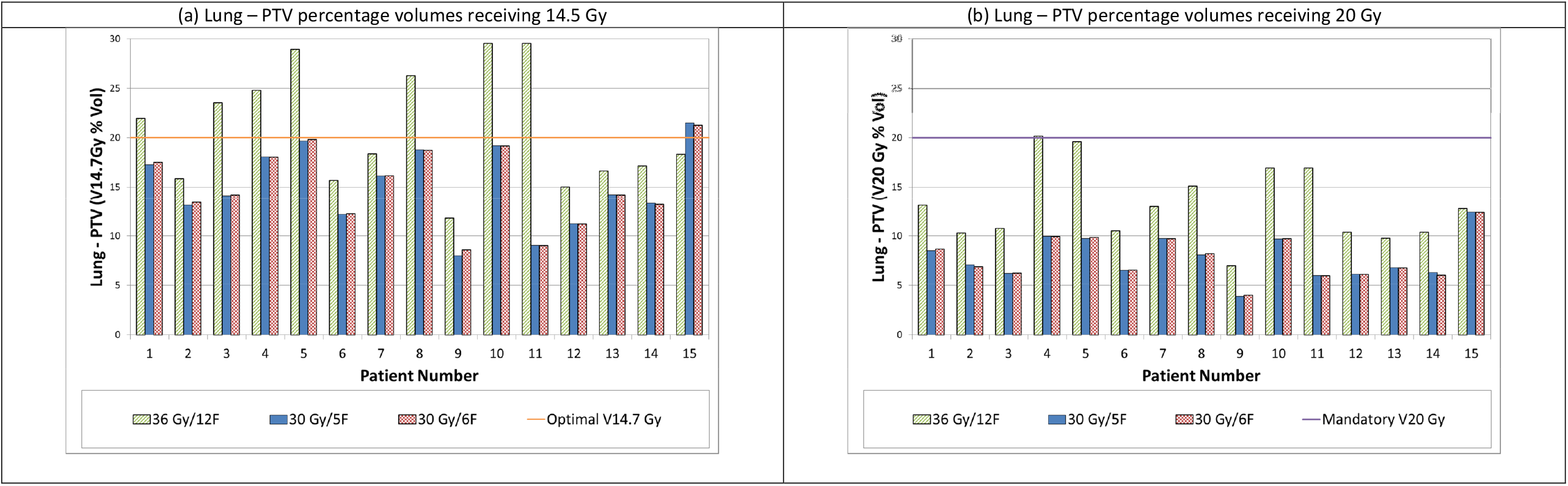
Lung – PTV percentage volumes receiving (a) 14.7 Gy and (b) 20 Gy for patients 1-15 and planned for 36 Gy in 12F (green striped), 30 Gy in 5F (solid blue) and 30 Gy in 6F (red hashed) respectively. The relevant mandatory (thick purple) and optimal (thin orange) constraints are indicated in terms of the BED converted from the dose constraint defined for 36 Gy/12F.

**Figure 3:**
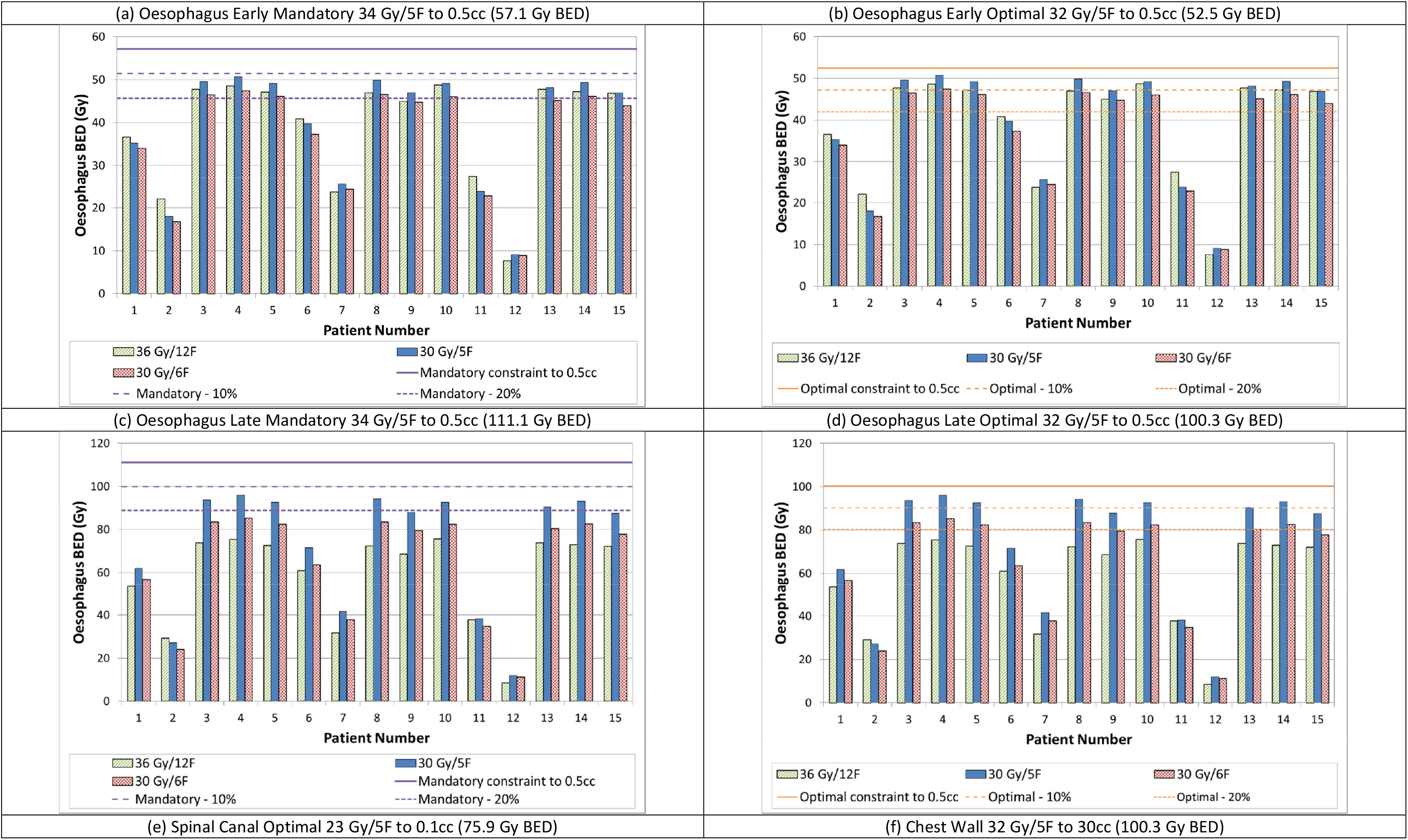
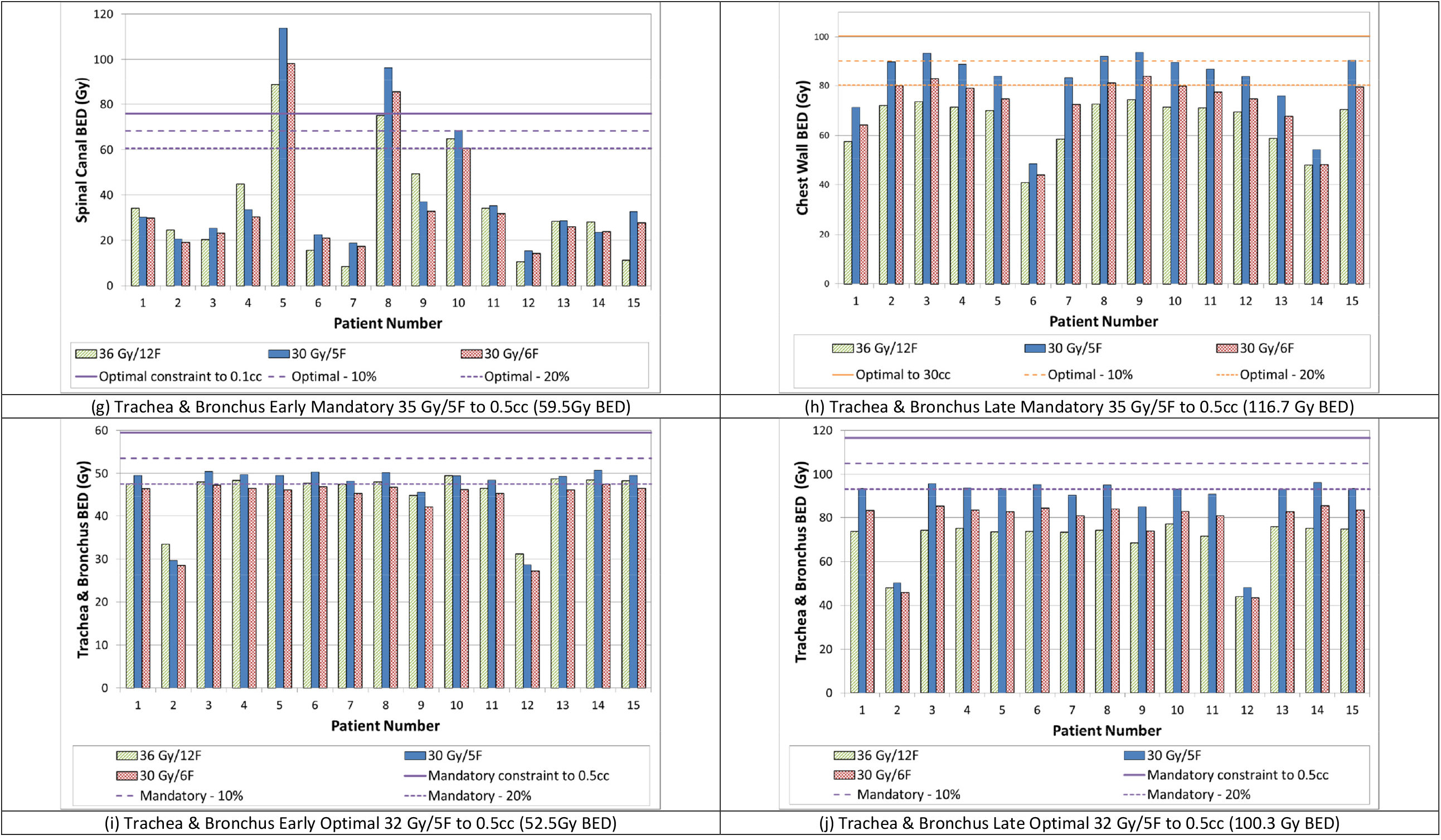
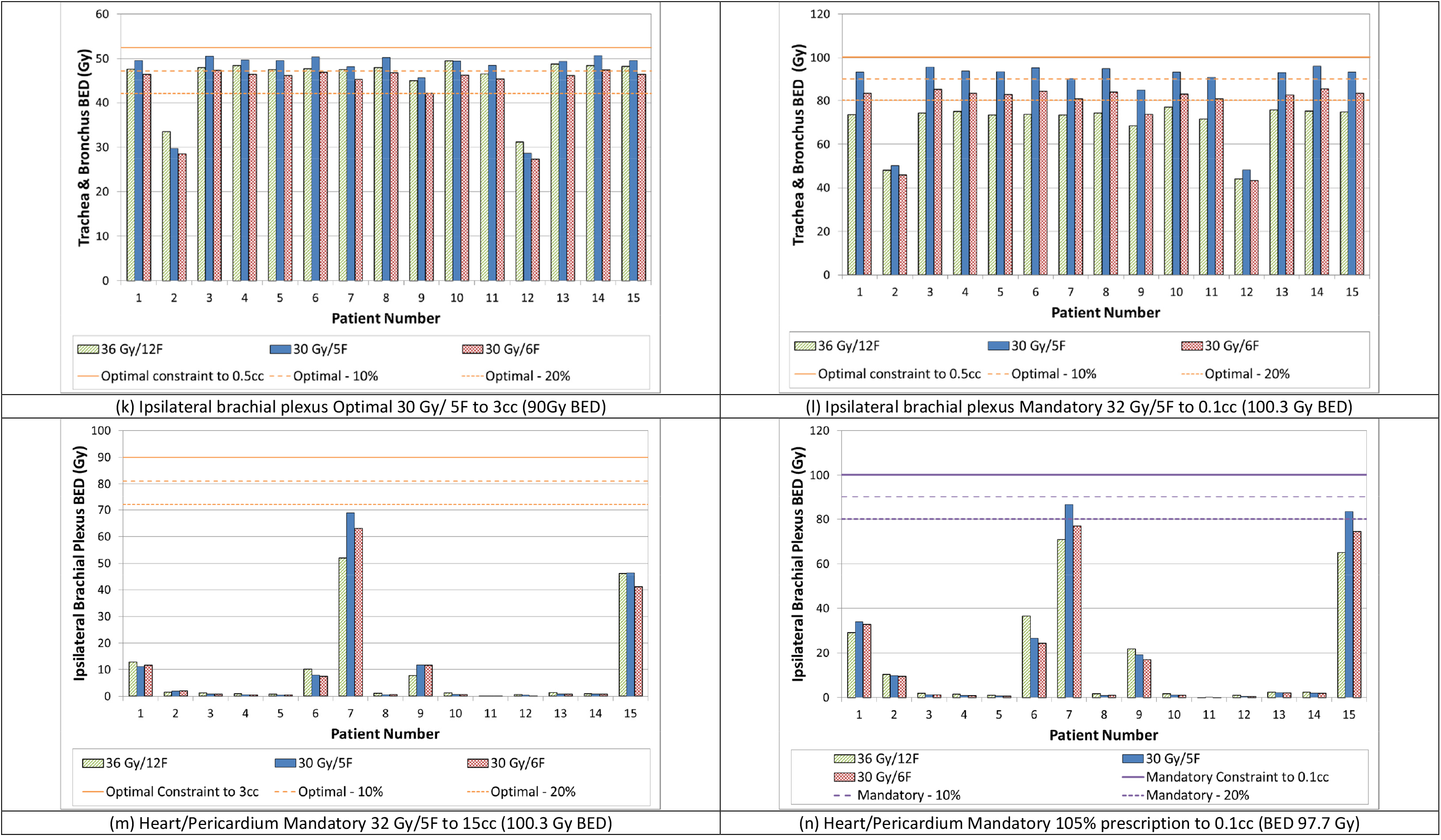
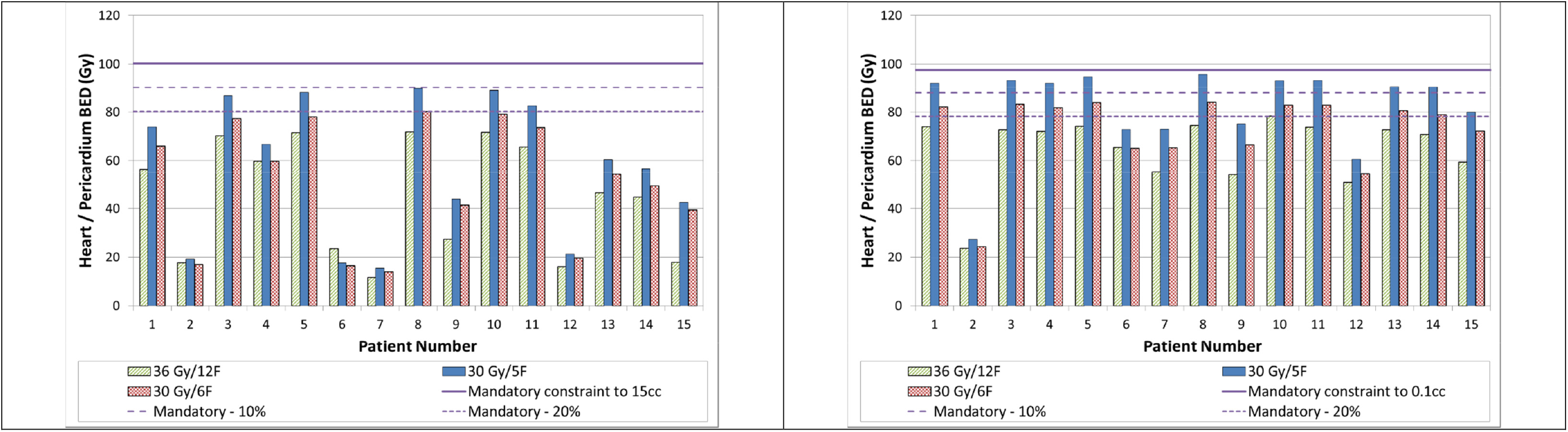
Biologically Equivalent Doses (BEDs) to various OARs for 15 patients (numbered 1 – 15). Fractionations of 36 Gy in 12F, 30 Gy in 5F and 30 Gy in 6F are shown as indicated by striped green, solid blue, and hashed red, respectively. The relevant mandatory (thick purple) and optimal (thin orange) constraints are indicated in terms of the BED converted from the dose constraint defined for 36 Gy/12F. Long and short dashed lines indicate the estimated relevant constraints with chemotherapy (standard constraint – 10%) and with chemotherapy plus surgery (standard constraint – 20%) respectively. Data are shown for (a) Oesophagus early mandatory 34 Gy/5F to 0.5cc (57.1 Gy BED), (b) Oesophagus early optimal 32 Gy/5F to 0.5cc (52.5 Gy BED), (c) Oesophagus late mandatory 34 Gy/5F to 0.5cc (111.1 Gy BED), (d) Oesophagus late optimal 32 Gy/5F to 0.5cc (100.3 Gy BED), (e) Spinal canal optimal 23 Gy/5F to 0.1cc (75.9 Gy BED), (f) Chest wall 32 Gy/5F to 30cc (100.3 Gy BED), (g) Trachea & Bronchus early mandatory 35 Gy/5F to 0.5cc (59.5Gy BED), (h) Trachea & Bronchus late mandatory 35 Gy/5F to 0.5cc (116.7 Gy BED), (i) Trachea & Bronchus early optimal constraint 32 Gy/5F to 0.5cc (52.5Gy BED), (j) Trachea & Bronchus late optimal constraint 32 Gy/5F to 0.5cc (100.3 Gy BED), (k) Ipsilateral brachial plexus optimal 30 Gy/ 5F to 3cc (90 Gy BED), (l) Ipsilateral brachial plexus mandatory 32 Gy/5F to 0.1cc (100.3 Gy BED), (m) Heart/Pericardium mandatory 32 Gy/5F to 15cc (100.3 Gy BED), (n) Heart/Pericardium mandatory 105% prescription to 0.1cc (BED 97.7 Gy

A summary of the PTV characteristics is given in Table 4 which includes the PTV volume and the location of the PTV relative to OARS. The three plans selected for treatment delivery verification all passed the standard tolerances that are used locally. This suggests that all the 5F and 6F plans produced would be dosimetrically accurate and clinically deliverable.

**Table 4:** Tumour locations with respect to OARs and the optimal dose constraints exceeded.

| **Plan** | **Tumour location** | **PTV Volume (cc)** | **Overlapped PTV & OAR** | **OAR <1 cm of PTV** | **Optimal dose constraints exceeded (5F)** | **Optimal dose constraints exceeded (6F)** |
| --- | --- | --- | --- | --- | --- | --- |
| 1 | Rt. Central | 354.82 | Heart | Oesophagus | - | - |
| 2 | Rt. Post. | 313.46 | - | - | - | - |
| 3 | Lt. Central | 441.82 | Heart<br>Oesophagus | - | Oesophagus | Oesophagus |
| 4 | Rt. Post. & Central | 482.85 | Heart<br>Oesophagus | - | Oesophagus | Oesophagus |
| 5 | Rt. Central / Post. | 358.84 | Heart<br>Oesophagus | - | Oesophagus<br>Spinal canal<br>PRV | Oesophagus<br>Spinal canal<br>PRV |
| 6 | Lt. Sup. | 256.87 | - | Oesophagus | - | - |
| 7 | Lt. Ant. | 200.65 | - | Oesophagus<br>Heart | - | - |
| 8 | Lt. Post. | 696.41 | Spinal Canal<br>PRV<br>Oesophagus<br>Heart | Spinal Canal<br>PRV | Oesophagus<br>Spinal canal<br>PRV | Oesophagus<br>Spinal canal<br>PRV |
| 9 | Rt. Post. | 541.38 | Oesophagus | Spinal Canal<br>PRV<br>Heart | - | - |
| 10 | Rt. Central / Post. | 759.06 | Heart,<br>Oesophagus | Spinal Canal<br>PRV | Oesophagus<br>Spinal canal<br>PRV | Oesophagus |
| 11 | Lt. Post. | 312.98 | Heart | Oesophagus<br>Spinal Canal<br>PRV | - | - |
| 12 | Rt. Peripheral | 251.85 | - | Heart | - | - |
| 13 | Rt. Central | 235.48 | Oesophagus<br>Heart | - | Oesophagus | - |
| 14 | Rt. Central | 235.10 | Oesophagus<br>Heart | - | Oesophagus | Oesophagus |
| 15 | Rt. Sup. / Central | 413.93 | Oesophagus | Heart | - | - |

## Discussion

It is well recognised that setting up large trials comparing different palliative thoracic RT regimens is difficult and is confounded by the increasing use of systemic chemotherapy (24). Trials aimed at integrating palliative radiotherapy and systemic therapy are needed, particularly for patients with good PS (24). Considerable care is necessary to avoid severe life changing side effects. For example, the studies of Dische *et al*., and Macbeth *et al*., show that RM can occur although that was probably associated with treatment planning limitations in earlier times (29,42). The study by Dische and colleagues is important since 6 fraction schedules were used and they retrospectively found that spinal cord doses over 33 Gy could cause RM although non-coplanar dose estimations were perhaps not as accurate as those 3-D dose distributions available now (29,42).

With more recent advances in radiotherapy planning, verification and delivery techniques, it may be possible to safely treat patients with adequate dose of palliative radiation in fewer fractions. It is possible to keep doses to normal tissues and organs within tolerance by carefully adhering to the established dose constraints during planning and robust verification before delivery. The current planning study suggests that it is probably safe to deliver high dose palliative radiotherapy to a similar BED without any expected significant increase in toxicity in the case of most patients. However, the study suggests that care must be taken. The spinal canal PRV dose constraints shown in Figure 1 (c) and (d) indicate that plans 5, 8 and 10 meet the mandatory constraint but exceed the conservative 100 Gy BED constraint as this was not included within the optimisation goals. This is unsurprising perhaps due to the position of all 3 tumours in the posterior part of the lung and in close proximity to the spine. Plans 8 and 10 are for large tumours, which may indicate that some selection of appropriate tumour size may be appropriate, above which this fractionation change is less tolerable. Seven of the fifteen plans did not meet the optimal oesophageal constraints (3, 4, 5, 8, 10, 13 and 14) and this rose to eight (Plan 15) if the constraint minus 20% cut-off was used. However, using the late mandatory oesophageal constraints, all plans pass with 30 Gy/ 6F. The plans which fail the early mandatory oesophageal constraints also would have failed these using the standard 36 Gy/12F, and the use of VMAT for the 30 Gy/6F reduced these doses. Normal lung (Lung – PTV) doses are also improved owing to the use of VMAT as seen in Figure 2, and all are below the mandatory constraint. Mandatory heart constraints were met in all cases for the 30 Gy/ 6F plans.

Out of the cohort of fifteen patients, 5F plans for all patients met all of the mandatory constraints when the full BED was used. This was reduced to six patients when the 10% reduction due to the chemotherapy is considered and further reduced to three patients when the 20% reduction is considered. When the tolerances are not fully met, or if the BEDs are close to the tolerance values, switching from 5F to 6F may reduce toxicity whilst still delivering comparable BEDs to 12F. The number of plans meeting all mandatory constraints for each fractionation is given in Table 5.

**Table 5:**
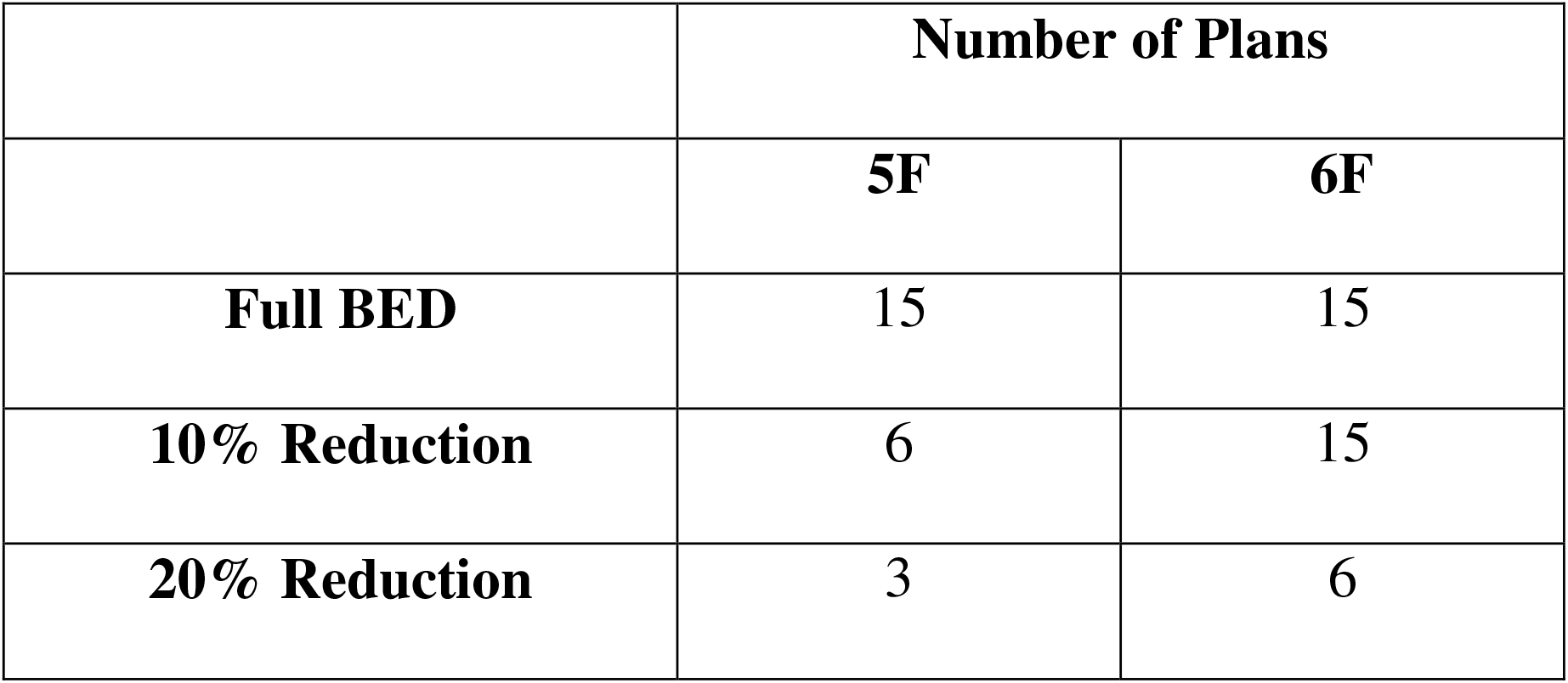
Number of 5F and 6F plans meeting all mandatory constraints for different BED scenarios.

Otherwise, if there are concerns, the total dose could be reduced to 27.5 Gy or lower, and/or further increased fractionation used as guided by BED estimations. Another intriguing possibility is to electively give the same dose-fraction but in a longer treatment session time in carefully selected patients, since such an approach will reduce the BED. For example, a change from 5-10 minutes to 20-25 minutes may be sufficient to reduce spinal cord BED significantly (34) since half of the sub-lethal damage repair is eliminated with a half-time of 12 minutes.

This needs to be confirmed in a phase II safety and feasibility study. The outcomes from such a study could potentially reduce both the number of patient hospital visits for radiotherapy and the overall duration of radiotherapy, thus benefiting the patient and the clinical service.

There are limitations, inherent biases and potential criticisms in any study. This is a planning study of patients already treated using the standard high dose palliative radiotherapy (36 Gy in 12 F). No assessment or comparison of clinical outcomes or toxicity was possible, but the BED method is the simplest way to estimate whether any iso-effective dose is exceeded.

The patients and the scans were selected in order ensure representation of diverse anatomical locations, staging, tumour sizes, and volumes and therefore it was not deemed appropriate to perform statistical analyses beyond the binary question of whether a safe and clinically usable radiotherapy plan could be generated for these dose-fractionation regimes. The dose-fractionation calculations are based on the linear quadratic model of radiation effect but has been used cautiously.

There is a potential health economic impact which would follow reduced patient attendances, allowing reduction of waiting lists for other treatments, especially those which involve single fractions or other palliative schedules. The proposed shorter regime would typically have at least 6 fewer fractions for every patient. By adopting the shorter dose-fractionation regime, there would be substantial cost-saving benefits for the NHS (year on year), resulting from fewer fractions of radiotherapy for each patient, to achieve similar outcomes. Furthermore, opportunity benefits from optimization of resource utilisation, by freeing-up some radiotherapy appointments (at least 6 fractions per patient), would consequently increase available treatment capacity, with potentially reduced radiotherapy waiting times for other patients.

## Conclusion

It has been demonstrated that it is potentially safe and feasible to deliver high dose palliative radiotherapy using the 5F or 6F regimes described, when planned to comparable target BEDs to the current 12F standard. It has also been suggested that the toxicity from either of these regimes would be within acceptable limits provided that the dose constraints described can be adhered to. However, careful patient selection may be required with respect to target volume size, target volume location and the potential impact of chemotherapy or other interventions.

A Phase II study would be required to fully assess the safety and feasibility of the 5F and 6F regimes. The outcomes from such a study could potentially reduce the number of patient hospital visits for radiotherapy, thus benefiting the patient and the clinical service by releasing treatment capacity therefore optimising resource utilisation.

## Ethics Approval

This is a retrospective planning study using anonymised patient CT data. GafREC approval was sought and given (UHCW GF0401) and full ethics approval was waived as study type is excluded from NHS Research Ethics Committee review.

## Data Availability

All data produced in the present study are available upon reasonable request to the authors.

